# STUNTING PREVENTION BASED ON HEALTH PROMOTION MODEL FROM PERSPECTIVE PHILOSOPHY OF SCIENCE: A LITERATURE REVIEW

**DOI:** 10.1101/2022.09.19.22280088

**Authors:** Hurun Ain, Moses Glorino Pandin, Yuni Sufyanti Arief

**Affiliations:** Doctor of Nursing, Faculty of Nursing Universitas Airlangga, Jalan Dr. Ir. H. Soekarno, Mulyorejo, Kec. Mulyorejo, Surabaya, East Java 60115; Faculty of Nursing Universitas Airlangga, Jalan Dr. Ir. H. Soekarno, Mulyorejo, Kec. Mulyorejo, Surabaya, East Java 60115; Faculty of Humanities Universitas Airlangga, Jalan Dr. Ir. H. Soekarno, Mulyorejo, Kec. Mulyorejo, Surabaya, East Java 60115; Health Polytechnic of the Ministry of Health Malang, Jalan Besar Ijen 77 C Malang Kec. Klojen Malang City, East Java

## Abstract

**Introduction:** Stunting is one of the biggest nutritional problems in toddlers in Indonesia which is based on Nutrition Status Monitoring (PSG) data for the last three years. The factor that has a large influence on the incidence of stunting is the pattern of parental care when the child is at the age of 0-59 months. The aim of this study is to study stunting prevention from the perspective of the theory of health promotion model

**Method:** This study uses a literature review using PICOS framework. Articles were searched from 3 databases, namely PubMed, Sciencedirect, Proquest. The keywords used in the literature search were: 1) ((Stunting prevention) AND (pender’s) OR (health promotion model), 2) Stunting prevention AND pender’s OR health promotion model; 3) stunting prevention. The search is limited to publications in 2018-2022, free full text, in English, not Reviews. Then selected using the PRISMA diagram and obtained 14 articles. Quality assessment of 14 articles using JBI Critical Appraisal tools.

**Result:** Seven articles stated that the determinant factor of stunting was the low education of parents (mothers) which resulted in poor parenting of children. Seven articles discussing the Health Promotion Model (HPM) theory can be used as a guide in providing education to improve health promotion.

**Conclusion:** good parenting behavior can be improved by increasing mother’s knowledge through providing education using HPM theory as a basis

## Introduction

Stunting is one of the biggest nutritional problems in toddlers in Indonesia were based on Nutrition Status Monitoring (PSG) data for the last three years, stunting has the highest prevalence compared to other nutritional problems such as undernutrition, thinness, and obesity (Kemenkes, 2018).. Stunting is a less problem chronic malnutrition caused by lack of nutritional intake for a long time, resulting in impaired growth in children, namely the child’s height is more low or short (stunted) than the standard age. Factors that have a big influence on the incidence of stunting is a parenting pattern when the child is at the age of 0-59 month. This poor parenting pattern is closely related to inadequate parenting the practice of feeding (*feeding practice*) in children which results inadequate nutritional intake, both in quantity and quality which lasts for a long time. Apart from the practice of giving eating, which is no less important are parenting practices in prevention infectious diseases such as immunization, preventing children from exposure to infection, these clean and healthy living behavior habits will also result in directly on the occurrence of stunting.

The bad impact that can be caused by short-term stunting is disruption of brain development, intelligence, impaired physical growth, and metabolic disorders in the body. Stunting can hinder children’s physical and mental development, is associated with an increased risk of illness and death as well as stunted growth of motor and mental abilities. Toddlers who experience stunting have a risk of decreased intellectual ability, productivity, and increased risk of degenerative diseases in the future

Accelerating the reduction of stunting in Toddlers is a government priority program as stated in the 2020-2024 National Mid-term Development Plan (RPJMN). Prevention and reduction stunting in Indonesia is not just a government affair. All elements of the nation must be involved and play an active role in fighting stunting in Indonesia. To achieve this target, innovation efforts are needed to reduce the number of stunting toddlers to 2.7% per *2024*. year especially mothers about correct parenting for children aged 0-59 months, therefore it is necessary to study through a literature review on childcare based on health promotion with the approach of parent-child interaction methods.

## Search

literature for the authenticity of this study using articles in English originating from 3 databases: PubMed, Sciencedirect, Proquest. The keywords used in the literature search were: 1) ((Stunting prevention) AND (pender’s) OR (health promotion model), 2) Stunting prevention AND pender’s OR health promotion model; 3) stunting prevention. The search is limited to publications in 2018-2022, free full text, in English, not reviews.

The strategy used to search for articles using the PICOS framework consists of:

**Figure 1.**
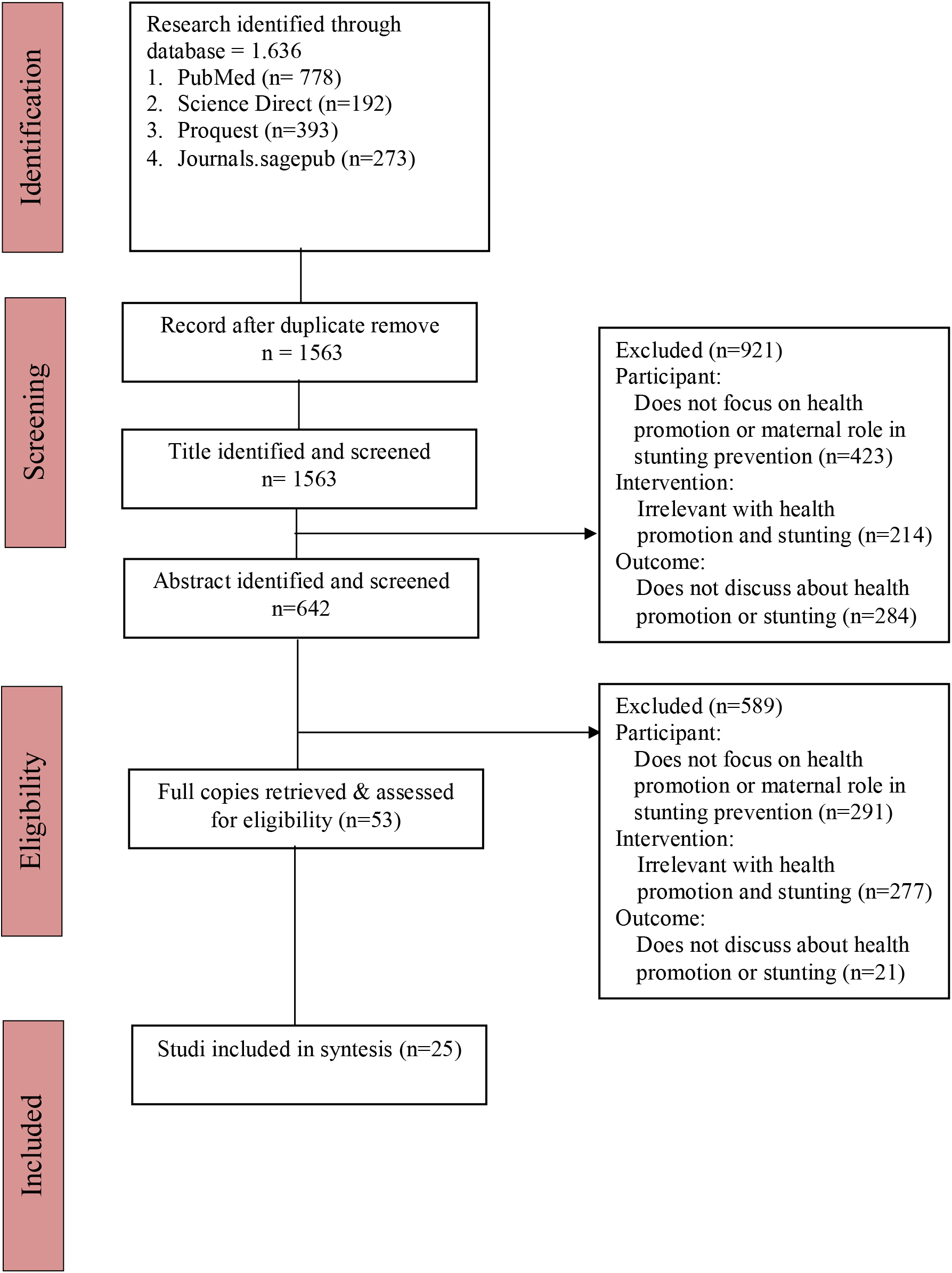
Literature Search Flow Diagram based on PRISMA

**Tabel 1.** Characteristic of Reviewed Studied

| No | Judul, Penulis, dan Tahun | Metode Penelitian | Hasil |
| --- | --- | --- | --- |
| 1. | **Ressa Andriyani Utami, *et al.*,**<br>Identifying causal risk factors for stunting in children under five years of age in South Jakarta, Indonesia, *Enfermería Clínica* 2019, Vol 29 page 606-611 | **Design:** cross-sectional study<br>**Sample:** 192<br>**Variables:**<br>Independent: family characteristics, nutritional parenting<br>Dependent: occurrence of stunting<br>**Instruments:**<br>**Analysis:** t-test, chi-square, and multiple logistic regression | There is significant correlation between family characteristics and nutritional parenting with the occurrence of stunting in children under five |
| 2. | **Faramarz Shaahmadi, *et al.*,**<br>Factors Influencing Health Promoting Behaviors in Women of Reproductive Age in Iran: Based on Pender's Health Promotion Model, *Open Access Macedonian Journal of Medical Sciences* 2019 Vol 7 (14), 2360-2364 | **Design:** cross-sectional study<br>**Sample:** 240 women aged 15-49 years<br>**Variables:** factors that influence health promotion behavior<br>based on the Pender model in women of childbearing age<br>**Instruments:** Questionnaire *health promoting lifestyle profile-II* (HPLP-II)<br>**Analysis:** descriptive, pearson correlation, t-test, ANOVA | There is a significant relationship significant between stress management and variables include *Perceived benefits of action, Perceived barriers to action, prior related behavior, situational influence, commitment to plan of action* |
| 3. | **Farideh Jalili Bahabadi, *et al.*,**<br>Predictors of Health Promoting Lifestyle in Pregnant Women Based on Pender's Health Promotion Model, *International Journal of Women's Health* 2020 Vol 12, 71-77 | **Design:** descriptive study<br>**Sample:** 300 pregnant women in trimester 2 and 3<br>**Variables:** lifestyle, construction of the HPM Pender model<br>**Instruments:** demographic questionnaire, Health Promoting Lifestyle Profile II (HPLP-II) and a questionnaire based on the construct of the Pender model<br>**Analysis:** Descriptive, | Among model constructs Pender, social support, perceived barriers, and the benefits felt to have significant influence to the lifestyle improve health |
|  |  | Pearson's correlation, multiple linear regression |  |
| 4. | **Rabiye Erenoglu, et al.,** The Effect of the Health Promotion Program for Young People on Health Behaviors, Health Perception, and Self Efficacy Levels: A Randomized Controlled Trial, International Journal of Caring Science, 2019, Vol 12 (2 ) | **Design:** randomized controlled experimental study<br>**Sample:** 37 students<br>**Variable:** Independent: health promotion program<br>Dependent: Health Behaviors, Health Perception, and Self Efficacy Levels<br>**Instrument:** Healthy Lifestyle Behavior Scale-II, the Perception of Health Scale, and the Self-Efficacy Scale<br>**Analysis:** X2 test, the Mann Whitney U Test, and Wilcoxon Signed Rank | There was a significant increase in the experimental group compared to the control group in terms of health responsibility, Healthy Living behavior, self-awareness, Health Perception |
| 5. | **Masoomeh Goodarzi Khoigan, et al.,** Impact of nutrition education in improving dietary pattern during pregnancy based on pender's health promotion model, Iranian Journal of Nursing and Midwifery Research 2018, Vol 23 (1) | **Design:** randomized clinical trial<br>**Sample:** 192 pregnant mothers<br>**Variable:** independent: nutrition education<br>intervention based on Pender's HPM<br>Dependent: dietary pattern<br>**Instrument:** Validated questionnaire<br>**Analysis:** 2 independents sample t tests, paired sample t test, Pearson correlation | Educational interventions based on the HPM pender model are effective in improving diet patterns in pregnant women |
| 6. | **Mille Hepburn,** The Variables Associated with Health Promotion Behaviors Among Urban Black Women, Journal of Nursing Scholarship 2018, Vol 50 (4) | **Design:** cross-sectional survey<br>**Sample:** 132 Urban Black women between the ages of 30 to 64<br>**Variables:** Variables Associated with Health Promotion Behaviors | health literacy, efficacy self, and readiness to change is related variables with promotional behavior health |
|  |  | **Instrument:** NVS, NGSE], HRI, HPLPII<br>**Analysis:** Univariate, linear regression |  |
| 7. | **Robabeh Haghi, et al.,** The role of correlated factors based on Pender health promotion model in brushing behavior in the 13–16 years old students of Guilan Iran, Italian Journal of Pediatrics 2021, 47 (1) | **Design:** cross-sectional study<br>**Sample:** 374 girls and 387 boys) at urban public schools<br>**Variable:** Independent: correlated based on the promotion model Pender's health<br>Dependent: tooth brushing behavior<br>**Instrument:** Validated questionnaire<br>**Analysis:** regression model | factors Perceived self-efficacy, situational influences, and perceived barrier to action has a significantly stronger relationship with commitment to plan of action |
| 8. | **Masumi Maehara, et al.,** Patterns and risk factors of double burden of malnutrition among adolescent girls and boys in Indonesia, PloS one 2019, Vol 14 (8) | **Design:** cross-sectional survey<br>**Sample:** 1,080 girls and 1,080 boys and 1,951 parents<br>**Variables:** nutritional status, socio-demographic characteristics, morbidity, dietary intake and physical activity and other relevant factors.<br>**Instruments:** Survey questionnaires, interviews, and assessments<br>**Analysis:** Multivariable logistic regression | Family type, socioeconomic status, adolescent education, maternal education are risk factors for the nutritional status of adolescent girls and boys in Indonesia |
| 9. | **Fantay Gebru, et al.,** Determinants of stunting among under-five children in Ethiopia: a multilevel mixed effects analysis of 2016 Ethiopian demographic and health survey data, BMC pediatrics 2019, Vol 19 (1) | **Design:** Health survey, Cross sectional study<br>**Sample:** 8855 children <5 years old, 640 community clusters<br>**Variables:** Independent: gender, birth weight, poor family, mother's education.<br>Dependent: stunting<br>**Instrument:** Ethiopian Demographic and Health Survey | Male gender, low birth weight, poor family, low maternal education are the determinants of stunting |

|  |  | **Analysis:** logistic regression |  |
| --- | --- | --- | --- |
| 10. | **Richard B Kajjura, et al.**, Effect of Nutrition Education on Knowledge, Complementary Feeding, and Hygiene Practices of Mothers with Moderate Acutely Malnourished Children in Uganda, Food and Nutrition Bulletin 2019, Vol 40 (2) | **Design:** cluster randomized controlled trial<br>**Design:** 438 mother–child pairs participating<br>**Variables:** Independent: maternal nutritional education and counseling<br>Dependent: prevalence of stunting<br>**Instruments:** Anthropometric measurements, SECA stadiometer<br>**Analysis:** 2, t tests, regression | Age, gender, vomiting, maternal height, early weaning related to child growth |
| 11. | **Febriana, Widya Rahmawati Nurhaeni, Nani**, Is There Any Relationship between Feeding Practices for Children Under Two Years of Age (6–23 Months) and Stunting? Comprehensive Child and Adolescent Nursing 2019, Vol 42 | **Design:** cross-sectional study **Sample:** 143 mothers with children aged 6-23 months<br>**Variables:** Independent: feeding practices, maternal age, maternal employment status, maternal educational level, newborn birthweight, sanitation and health behaviors, and household income<br>Dependent: stunting<br>**Instrument:** Questionnaire<br>**Analysis:** Mann–Whitney test, Kolmogorov–Smirnov test, and chi-square test | No relationship meaning between practice baby feeding and toddlers with stunting, there are maternal age relationship, status mother's job, level maternal education, LBW, sanitation, behavior health, and income household with stunting |
| 12. | **Sadaf Khan, et al.**, Determinants of stunting, underweight and wasting among children, BMC public health 2019, Vol 19 (1) | **Design:** Survey<br>**Sample:** 3071 Pakistani children aged 0-59 months<br>**Variables:** Mother's education level, stature, birth weight, maternal BMI, ANC, age at marriage,<br>**Instrument:** Questionnaire, anthropometry<br>**Analysis:** binary logistic | Low maternal education level, short stature, LBW, maternal BMI associated with stunting |

|  |  | regressions |  |
| --- | --- | --- | --- |
| 13. | **Sasongko, *et al.*,**<br>Determinants of<br>adolescent shortness<br>in Tanjungsari, West<br>Java, Indonesia,<br>Asia Pacific journal of<br>clinical nutrition 2019,<br>Vol 28 (suppl 1) | **Design:** cohort study<br>**Sample:** 3093 participating<br>children, their parents, and<br>households<br>**Variables:** birth weight, TB of<br>children aged 0-2 years,<br>mother's education, drinking<br>water sources, latrines,<br>exposure to infection at the age<br>of 0-2 years, exposure to<br>cigarette smoke, | Determinant<br>factors for short<br>adolescents: birth<br>of low birth weight,<br>short height at age<br>0-2 years,<br>inadequate<br>maternal education,<br>inadequate<br>drinking water<br>sources, B clock an<br>unfit, |
| 14. | **Hosseini**<br>**Khaboshan, *et al.*,**<br>Effect of a theory<br>based educational<br>intervention for<br>enhancing nutrition<br>and physical activity<br>among Iranian<br>women: a<br>randomized control<br>trial, public health<br>nutrition 2021, Vol 24<br>(18) | **Design:** randomized<br>controlled trial<br>**Sample:** 202 girls aged 4-6<br>years (102 treatment group,<br>100 control group)<br>**Variable:** Independent:<br>HPM Dependent:<br>nutritional status and<br>physical activity<br>**Instruments:** semi-<br>quantitative FFQ, Physical<br>Activity Questionnaire<br>(IPAQ-L)<br>**Analysis:** descriptive, t-test, chi<br>square, Pearson's correlations<br>ANOVA | Stunting increased<br>significantly in:<br>Children living in<br>families with<br>toddlers > 3<br>people; Family<br>members >6<br>people; ANC <4<br>times during<br>pregnancy; LBW<br><2,500 grams;<br>Decreased family<br>income |

## Results and Discussion

The results of the review of 14 articles found 2 articles explain about the determinants of stunting, namely research (Utami, Setiawan and Fitriyani, 2019) and (Sasongko *et al*., 2019), 5 articles discussing the relationship between parental/mother education and the incidence of stunting, namely research (Khan, Zaheer and Safdar, 2019), (Fantay Gebru *et al*., 2019), (Maehara *et al*., 2019). (Kajjura, Veldman and Kassier, 2019), and (Febriana and Nurhaeni, 2019). Articles discussing the application of the theory of Nola J. Pender *health promotion model* obtained 7 articles, namely his research (Shaahmadi *et al*., 2019), (Bahabadi *et al*., 2020), (Erenoglu, Rabiye, PhD; Can, Rana, PhD; Sekerci, Yasemin Gumus, 2019), (Goodarzi-Khoigani *et al*., 2018), (Hepburn, 2018), (Haghi *et al*., 2021), dan (Hosseini Khaboshan *et al*., 2021)

The study (Utami, Setiawan and Fitriyani, 2019), the results of parenting nutrition and family characteristics were found to be related to the incidence of stunting in children under the age of 5 years. The result of study (Sasongko *et al*., 2019) found that the determinants of short adolescents were: low birth weight birth, short height at the age of 0-2 years, inadequate maternal education, inadequate drinking water sources, inadequate latrines. The result of study (Khan, Zaheer and Safdar, 2019) shows that low maternal education levels, short stature, low birth weight, maternal BMI are associated with stunting. Stunting research (Fantay Gebru *et al*., 2019) shows that male sex, low birth weight, poor families, low maternal education are the determinants of stunting. Study was conducted by (Maehara *et al*., 2019) found that family type, socioeconomic status, adolescent education, maternal education are risk factors for the nutritional status of adolescent girls and boys in Indonesia. Study by (Kajjura, Veldman and Kassier, 2019) increasing age, gender, vomiting, maternal height, early weaning is associated with child growth. Study of (Febriana and Nurhaeni, 2019) shows that there is a relationship between maternal age, mother’s employment status, mother’s education level, low birth weight, sanitation, health behavior, and household income with stunting.

Mother’s education has a very large contribution to children’s health status, especially child development. Mothers who do not have adequate education and knowledge about good and correct parenting are not able to provide nutritional intake that is in accordance with the needs of children both in terms of quantity and quality. The inability of families to provide optimal nutritional intake in this era, especially in developing countries, is not caused by economic incapacity but is more often caused by inadequate parental knowledge about nutritional parenting and stimulation of child growth and development. Mother’s knowledge is closely related to the level of education, as a review conducted by Martorell 2012 in Phiri 2014 At least 25 studies show the level of mother’s education also affects the child’s stunting status (Phiri, 2014).

Poor parenting practices, including lack of knowledge of mothers about health and nutrition before and after during pregnancy and after delivery. Some facts and available information show that 60% of children aged 0-6 months do not receive exclusive breastfeeding, and 2 out of 3 children aged 0-24 months do not receive complementary feeding. Complementary feeding is given/started to be introduced when toddlers are over 6 months old. In addition to functioning to introduce new types of food to babies, complementary feeding can also meet the nutritional needs of the baby’s body which can no longer be supported by breast milk, as well as form the immune system and development of the child’s immunological system to food and drink (TNP2K, 2017)

The above is in line with the results of research (Sofiatin *et al*., 2019) and (Izwardy, 2019) which state that the occurrence of stunting is closely related to poor parenting behavior due to low maternal education. Mothers do not give exclusive breastfeeding, do not give proper complementary foods according to age, do not fully immunize their children, do not maintain sanitation of drinking water and the environment, do not take advantage of existing health facilities (Sofiatin *et al*., 2019)

Not implementing early initiation of breastfeeding, the failure of exclusive breastfeeding, and the early weaning process can be one of the factors for stunting. Meanwhile, in terms of providing complementary feeding things that need to be considered are the quantity, quality, and safety of the food provided (Kementerian Kesehatan RI, 2018). Problems with child growth will occur if continuous breastfeeding is not accompanied by adequate complementary feeding at the appropriate age. With increasing nutritional needs, if a child receives inadequate complementary foods, impaired linear growth may occur (Derso *et al*., 2017). Lestari, Hasanah, and Nugroho (2018) in their research concluded that stunting has a statistically significant relationship with the ASI variable. This study found that exclusive breastfeeding can reduce the prevalence of stunting in children under five (Suhariyanto, Muis and Et.al, 2020)

Research conducted by Semba, RD et al. in Indonesia found that children who do not receive immunizations are more at risk for malnutrition (malnutrition) and anemia and have higher rates of infectious disease morbidity. These unimmunized children are also at high risk of acute malnutrition (Suhariyanto, Muis and Et.al, 2020)

Infectious diseases caused by poor hygiene and sanitation (eg diarrhea and worms) can interfere with the absorption of nutrients in the digestive process. Some infectious diseases suffered by the baby can cause the baby’s weight to drop. If this condition occurs for a long time and is not accompanied by adequate intake for the healing process, it can result in stunting (Kementerian Kesehatan RI, 2018). The increased exposure to various diseases and conditions of children because of increasing age, such as exposure to poor food hygiene and environmental sanitation, may contribute to poor growth (Akombi *et al*., 2017). Infectious diseases are related to the high incidence of infectious diseases, especially diarrhea, intestinal worms, and acute respiratory diseases (ARI). Many of these factors are related to the quality of basic health services, especially immunization, environmental quality and healthy living behavior. The quality of the environment is mainly the availability of clean water, sanitation facilities and healthy living behaviors such as the habit of washing hands with soap, defecating in the latrine, not smoking, air circulation in the house and so on (Abbas and Haryati, 2021)

Inadequate education and knowledge of mothers causes mothers to not utilize health facilities optimally, especially those related to reproductive health, pregnancy management, pregnancy checks, child development checks, and treatment of children if they are sick. Health services are still limited, including ANC-Ante Natal Care (health services for mothers during pregnancy), post-natal care and quality early learning. Information gathered from the publications of the Ministry of Health and the World Bank states that the attendance rate of children at “Posyandu” has decreased from 79% in 2007 to 64% in 2013 and children have not had adequate access to immunization services. Another fact is that 2 out of 3 pregnant women have not consumed adequate iron supplements and there is still limited access to quality early learning services (only 1 in 3 children aged 3-6 years has not been registered in Early Childhood Education services) (TNP2K, 2017). The results of the research of Titaley *et al*., 2013 found that the likelihood of stunting in Indonesia increased significantly in children whose mothers during pregnancy visited the ANC less than four times (Titaley *et al*., 2019)..

The results of a review of 5 articles discussing the theory of *health promotion models* are as follows: research (Shaahmadi *et al*., 2019) states that there is a significant relationship between stress management and variables including *Perceived benefits of action, Perceived barriers to action, prior related behavior, situational influence, commitment to plan of action*. The results of the study (Bahabadi *et al*., 2020) concluded that among the constructions of the Pender model, social support, perceived barriers, and perceived benefits had a significant influence on a health-promoting lifestyle. The results of the study (Erenoglu, Rabiye, PhD; Can, Rana, PhD; Sekerci, Yasemin Gumus, 2019) stated that there was a significant increase in the experimental group compared to the control group in terms of health responsibility, healthy living behavior, self-awareness, health perception, while research (Goodarzi-Khoigani *et al*., 2018) concluded that educational interventions based on the pender HPM model were effective in improving diet patterns in pregnant women. Research (Hepburn, 2018) literation of health, self-efficacy, and readiness to change are variables related to health promotion behavior. Research (Haghi *et al*., 2021) shows that perceived self-efficacy, situational influences, and perceived barriers to action have a significantly stronger relationship with commitment to plan of action. Research (Hosseini Khaboshan *et al*., 2021) in the experimental group, the intervention program had a significant effect on all HPM constructs which included behavior *outcome, and the estimates for prior behaviours, self-efficacy, interpersonal influences, feeling, perceived benefits and barriers, commitment, and behavior outcomes*

The HPM model will affect knowledge about health in the community. Adequate knowledge will influence the mother’s behavior in parenting practices which in turn can prevent the risk of stunting in children. HPM is one way to describe human interaction with the physical and interpersonal environment in various dimensions. Health Promotion Model is one of the theoretical models used in the field of behavior change. Knowledge is a very important domain for the formation of one’s behavior. Central to HPM is Albert Bandura’s theory of social learning, which mentions the importance of cognitive processes in behavior change. HPM is similar in the construction of health belief models but is not limited to disease prevention behavior (Martha Raile Alligood, 2014).

The center of HPM is Albert Bandura’s social learning theory, which mentions the importance of cognitive processes in behavior change. Social learning theory, now entitled social cognitive theory, covers self-confidence: self-attribution, self-evaluation, and *self-efficacy. Self-efficacy* is the main construct of HPM. In addition, the value expectation model of human motivation that Feather (1982) describes, which supports rational and economic behavior, is important for model development. HPM is similar in construction to the health belief model (HBM) but HPM is not limited to explaining disease prevention behavior. HPM differs from the concept of the health belief model in that HPM does not include fear and threats as a source of motivation for health behavior. From this explanation, HPM has evolved to include behaviors to improve health and have the potential to last throughout life (Martha Raile Alligood, 2014)

## Conclusion

Based on the results of a review of 14 articles that have discussed stunting prevention from the perspective of the health promotion model, it can be concluded that the determinant factor of stunting is the low level of parental education that causes poor parenting practices. In addition, the HPM model can be applied in preventing stunting through the provision of health education to improve parenting practices for children and the result is stunting can be prevented

## Data Availability

All data produced in the present work are contained in the manuscript

https://drive.google.com/drive/u/0/my-drive

## Notes

### Competing Interest Statement

The authors have declared no competing interest.

### Clinical Protocols

https://drive.google.com/drive/u/0/my-drive

### Funding Statement

This study did not receive any funding

